# Genomic epidemiology of the SARS-CoV-2 epidemic in Zimbabwe: Role of international travel and regional migration in spread

**DOI:** 10.1101/2021.01.04.20232520

**Authors:** Tapfumanei Mashe, Faustinos Tatenda Takawira, Leonardo de Oliveira Martins, Muchaneta Gudza-Mugabe, Joconiah Chirenda, Manes Munyanyi, Blessmore V Chaibva, Andrew Tarupiwa, Hlanai Gumbo, Agnes Juru, Charles Nyagupe, Isaac Phiri, Portia Manangazira, Alexander Goredema, Sydney Danda, Israel Chabata, Janet Jonga, Rutendo Munharira, Kudzai Masunda, Innocent Mukeredzi, Douglas Mangwanya, The COVID-19 Genomics UK (COG-UK) Consortium, SARS-CoV-2 Research Group, Alex Trotter, Thanh Le Viet, Gemma Kay, David Baker, Gaetan Thilliez, Ana-Victoria Gutierrez, Justin O’Grady, Maxwell Hove, Sekesai Mutapuri-Zinyowera, Andrew J. Page, Robert A. Kingsley, Gibson Mhlanga

**Affiliations:** National Microbiology Reference Laboratory, Ministry of Health and Child Care, Zimbabwe, P.O. Box ST 749, Southerton, Harare, Zimbabwe; Quadram Institute Bioscience, Norwich, United Kingdom; World Health Organization, Zimbabwe, 82-84 Enterprise Road, Highlands P.O. Box CY 348 Causeway, Harare, Zimbabwe; Department of Surgical Sciences, Faculty of Medicine and Health Sciences, University of Zimbabwe; Directorate of Pharmacy Services, Ministry of Health and Child Care, Zimbabwe; Ministry of Health and Child Care, Zimbabwe; Harare City Health Department, Beatrice Road Infectious Diseases Hospital, Harare, Zimbabwe; School of Biological Sciences, University of East Anglia, Norwich, United Kingdom; SARS-CoV-2 Research Group

**Keywords:** sequencing, virus, transmission, epidemiology, COVID-19

## Abstract

Zimbabwe reported its first case of SARS-Cov-2 infection in March 2020, and case numbers increased to more than 8,099 to 16th October 2020. An understanding of the SARS-Cov-2 outbreak in Zimbabwe will assist in the implementation of effective public health interventions to control transmission. Nasopharyngeal samples from 92,299 suspected and confirmed COVID-19 cases reported in Zimbabwe between 20 March and 16 October 2020 were obtained. Available demographic data associated with those cases identified as positive (8,099) were analysed to describe the national breakdown of positive cases over time in more detail (geographical location, sex, age and travel history). The whole genome sequence (WGS) of one hundred SARS-CoV-2-positive samples from the first 120 days of the epidemic in Zimbabwe was determined to identify their relationship to one another and WGS from global samples. Overall, a greater proportion of infections were in males (55.5%) than females (44.85%), although in older age groups more females were affected than males. Most COVID-19 cases (57 %) were in the 20-40 age group. Eight lineages, from at least 25 separate introductions into the region were found using comparative genomics. Of these, 95% had the D614G mutation on the spike protein which was associated with higher transmissibility than the ancestral strain. Early introductions and spread of SARS-CoV-2 were predominantly associated with genomes common in Europe and the United States of America (USA), and few common in Asia at this time. As the pandemic evolved, travel-associated cases from South Africa and other neighbouring countries were also recorded. Transmission within quarantine centres occurred when travelling nationals returning to Zimbabwe. International and regional migration followed by local transmission were identified as accounting for the development of the SARS-CoV-2 epidemic in Zimbabwe. Based on this, rapid implementation of public health interventions are critical to reduce local transmission of SARS-CoV-2. Impact of the predominant G614 strain on severity of symptoms in COVID-19 cases needs further investigation.

## Introduction

‘Severe Acute Respiratory Syndrome Coronavirus 2’ (SARS-CoV-2) was confirmed as the causative agent of ‘Coronavirus Disease 2019’ (COVID-19) (Huang *et al*., 2020). The virus spread to more than 200 countries, led to more than 47 million confirmed infections and more than 1.2 million deaths (as of 4^th^ of November 2020). The COVID-19 pandemic constitutes the largest global public health crisis in a century, with daunting health and socioeconomic challenges (Khandelwal, 2020). Countries in the African continent have particular challenges with respect to controlling COVID-19 infections and reducing mortality rates; coupled with weak public health delivery systems they have large populations of vulnerable people e.g. those immunocompromised from HIV/AIDS, with non-communicable diseases such as diabetes, hypertension and anaemia, or malnutrition (Kaseje, 2020).

The first COVID-19 case in Africa was recorded in Egypt on the 14th of February, 2020 (Lone and Ahmad, 2020), and on 27^th^ February, the sub-Saharan African region recorded its first case in Nigeria. Further cases in sub-Saharan Africa were recorded in South Africa and Zimbabwe, on March 5^th^ and 20^th^, respectively (Msomi *et al*., 2020). The initial cases identified in Africa were mostly introduced from Europe, the Middle East and the United States of America (Hâncean *et al*., 2020; Lone and Ahmad, 2020; Mehtar *et al*., 2020).

On 30^th^ March, after confirmation of the first death (24^th^ March 2020) Zimbabwe introduced a 21-day self-quarantine for citizens arriving from affected territories. On the 23rd of March 2020, the country banned: non-essential in-bound travel and transportation except for returning residents; entertainment and recreational activities; public gatherings of > 50 people; and limited hospital visits to one relative per patient per day. Other measures implemented included compulsory wearing of face masks in all settings, social distancing and hand sanitization. A national COVID-19 coordinating committee chaired by a Cabinet Minister, with a full time senior public health specialist as its coordinator, was set up. At all levels from province to district, coordinating committees were established. Rapid Response Teams at all levels were set up to investigate presumed COVID-19 cases, follow-up on all contacts, collect samples from contacts and supervise the burial of confirmed COVID-19 deaths. Treatment centres were set up, mainly Harare and Bulawayo city. Due to the lockdowns imposed in other countries, there was an influx of Zimbabwean residents returning home. This led to establishment of Government run quarantine centres. Initially, all cases housed at quarantine centres had follow up laboratory testing on day 1, 7 and 21 to assess infection and recovery rate. After having completed the 21-day quarantine period with negative results, the returnees/travellers were discharged. The intervention meant that even those who were not showing signs and symptoms of COVID-19 were tested. On 30^th^ March, total shutdown of all services, except essential services like health care centres, grocery stores and government activities was implemented.

The COVID-19 pandemic, and the actions taken in response to it, disrupted healthcare services and, as a result, overwhelmed the major healthcare programmes in Africa focused on controlling HIV, tuberculosis and malaria. Facilities and staff are taken up with attending to the overwhelmingly high demand for care required by patients with severe COVID-19, leaving other patients without the resources they need. This will have far-reaching consequences for other diseases, poverty, food security, and economic growth in Africa (Walker *et al*., 2020).

There is an urgent need to understand the epidemiology of SARS-CoV-2 in Africa, particularly the dynamics of its virulence, transmission, mutation capacity and symptom variation amongst infected people (Kalk and Schultz, 2020). The SARS-CoV-2 mutation rate is estimated to be approximately 2.5 single nucleotide polymorphisms (SNPs) per month (Duchene *et al*., 2020; Meredith *et al*., 2020). Genome sequencing of SARS-CoV-2 is important to monitor these changes because they are relevant to vaccine efficacy, can be used to infer transmission dynamics, and inform future public health measures targeting its spread. Genomic surveillance backed up with detailed epidemiological data on SARS-CoV-2 can identifies which lineages of the virus are circulating in the human population. Information on how these change over time can serve as an indicator of the success of control measures, how often new sources of virus are introduced from other geographical areas, and how the virus has evolved in response to interventions. Despite having inadequate health systems, the number of reported cases in many African countries, specifically Zimbabwe, has been rising more slowly than predicted (Nkengasong and Mankoula, 2020).

Zimbabwe took the initiative to sequence SARS-CoV-2 samples from 100 patients collected over the period March to June 2020. The objectives were to use the sequence results to: understand initial transmission events; understand early domestic transmission of the virus in Zimbabwe; add context to the regional and global data; and to evaluate the role of rapid whole genome sequencing for outbreak analysis in this setting.

## Material and methods

### Epidemiological analysis

Nasopharyngeal samples (92,299 in total including repeats) were collected from suspected COVID-19 cases across Zimbabwe between the 20^th^ March (first case identified) and the 16^th^ October 2020 were diagnosed routinely at nine local laboratories to various regions affected using previously described PCR methods to confirm positive cases (Corman *et al*., 2020). Additional laboratories were established during the national scale up to help management of COVID-19 pandemic. At sampling the demographic data including gender, age and history of travel were recorded. Demographic, clinical and laboratory data associated with the positive cases (8,099) was collated and analysed in order to describe the epidemiology of SARS-Cov-2 using an analytical cross-sectional study design. Specifically, we determined the number of SARS-CoV-2 infections detected on each day since the first case in Zimbabwe, the distribution of detected cases in ten geographical regions of Zimbabwe, the number of infections in males and females, and the number of infection by age stratification. Access to these data (and associated clinical samples used for genomic analysis) was through coordination with teams from the Epidemiology and Diseases Control Unit of the Ministry of Health and Child Care of Zimbabwe.

### Selection of samples for genomic evaluation

Samples from the first cases (244) by date of collection (20th March to 30th June) that gave a positive PCR test were considered for genome sequencing. Samples that had a Ct value of less than 30 in the original PCR test were processed for sequencing as described below. Of these, 100 samples provided good quality sequence (<30% N’s) were used analysed by genome sequence comparison and phylogenetic analysis.

### RNA extraction

Samples for whole genome sequencing were processed using the QuantStudio 3 (Applied Biosystems) diagnostic platform at the National Microbiology Reference Laboratory (NMRL), Zimbabwe. RNA was extracted from nasopharyngeal swab samples using the NucliESNS easyMag (Biomerieux) instrument according to the manufacturer’s instructions. Left over aliquots of the RNA used for realtime PCR detection were transported to UK for sequencing. The lower cycle threshold (Ct) or take-off value produced by the SARS-CoV-2 assays in the QuantStudio 3 assays were used to determine whether samples needed to be diluted according to the ARTIC protocol (Quick, 2020).

### ARTIC SARS-CoV-2 multiplex tiling PCR

cDNA and multiplex PCR reactions were prepared following the ARTIC nCoV-2019 sequencing protocol v2 (Quick, 2020). Dilutions of RNA were prepared when required based on Ct values following the guidelines from the ARTIC protocol. V3 CoV-2 primers (https://github.com/artic-network/articncov2019/tree/master/primer_schemes/nCoV-2019/V3) were used to perform the multiplex PCR for SARS-CoV-2 according to the ARTIC protocol (Quick, 2020) with minor changes. Due to variable Ct values all samples in the two ARTIC multiplex PCRs were run for 35 cycles. Odd and even PCR reactions were pooled and cleaned using a 1x SPRI bead clean using KAPA Pure Beads (Roche Catalogue No. 07983298001) according to manufacturer instructions. PCR products were eluted in 30 µl of 10 mM Tris-HCL buffer, pH 7.5. cDNA was quantified using QuantiFluor ® ONE dsDNA System (Promega, WI, USA). Libraries were then prepared for sequencing on the Illumina or Nanopore platform and sequenced as described previously (Baker *et al*., 2020). All sequence data are freely available in the GISAID database (Supplementary Table 1 and Supplementary Table 2). Sequence data of SARS-CoV-2 from South Africa was reported previously (Msomi *et al*., 2020).

**Table 1.** SARS-CoV-2 cases and deaths per province (as of 16^th^ October 2020)

| Province | Population (est. 2019) | Total number of cases | Infection rate (per 100,000 population) | Number of deaths | Case Fatality Rate (CFR) |
| --- | --- | --- | --- | --- | --- |
| Bulawayo | 700,466 | 1,545 | 221 | 47 | 3.0 |
| Harare | 2,276,285 | 3,286 | 144 | 121 | 3.7 |
| Manicaland | 1,879,129 | 499 | 27 | 22 | 4.4 |
| Mashonaland Central | 1,235,657 | 208 | 17 | 4 | 1.9 |
| Mashonaland East | 1,441,974 | 412 | 29 | 4 | 1.0 |
| Mashonaland West | 1,609,978 | 350 | 22 | 12 | 3.4 |
| Midlands | 1,731,435 | 644 | 37 | 9 | 1.4 |
| Masvingo | 1,592,217 | 236 | 15 | 2 | 0.8 |
| Matabeleland North | 803,048 | 141 | 18 | 3 | 2.1 |
| Matabeleland South | 733,226 | 778 | 106 | 7 | 0.9 |
| **Total** | **14,003,415** | **8,099** | **58** | **231** | **2.9** |

**Table 2:** Summary of global lineages circulating in Zimbabwe, South Africa and Globally (as of 2020-11-08).

| Lineage | Zimbabwe (n=100) |  | South Africa (n=420) |  | Global (n=62138) |  |
| --- | --- | --- | --- | --- | --- | --- |
|  | no. samples | Percent | no. samples | Percent | no. samples | Percent |
| A | 4 | 4 | 1 | 0.08 | 711 | 0.5 |
| B.1 | 25 | 25 | 61 | 4.98 | 21383 | 15 |
| B.1.1 | 51 | 51 | 274 | 22.37 | 31648 | 22.1 |
| B.1.5 | 9 | 9 | 45 | 3.67 | 6253 | 4.4 |
| B.1.23 | 6 | 6 | 0 | 0 | 12 | 0.01 |
| B.1.1.57 | 3 | 3 | 20 | 1.63 | 22 | 0.02 |
| B.1.1.62 | 1 | 1 | 18 | 1.47 | 24 | 0.02 |
| B.2 | 1 | 1 | 1 | 0.08 | 2083 | 1.5 |

### Sequence analysis

The raw reads were demultiplexed using bcl2fastq (v2.20) (Illumina Inc.) allowing for zero mismatches in the dual barcodes to produce FASTQ files. The reads were used to generate a consensus sequence for each sample using an open source pipeline adapted from https://github.com/connor-lab/ncov2019-artic-nf (https://github.com/quadram-institute-bioscience/ncov2019-artic-nf/tree/qib). Briefly, the reads had adapters trimmed with TrimGalore (https://github.com/FelixKrueger/TrimGalore), were aligned to the Wuhan Hu-1 reference genome (accession MN908947.3) using BWA-MEM (v0.7.17) (Li, 2013), the ARTIC amplicons were trimmed and a consensus built using iVAR (v.1.2) (Grubaugh *et al*., 2019).

### Quality control of the sequenced genomes

Samples were prepared and sequenced in 96-well plates with one cDNA negative control per plate and one RNA extraction negative control, where applicable. Contaminated samples were removed from analysis. Consensus sequence was defined as passing quality control if greater than 50% of the genome was covered by confident calls or there was at least one contiguous sequence of more than 10,000 bases and no evidence of contamination in the negative control. This is regarded as the minimum amount of data to be phylogenetically useful. A confident call is defined as having 10x depth of coverage. If the coverage falls below these thresholds, the bases are masked with Ns. Low quality variants are also masked with Ns. The QC threshold for inclusion in Global Initiative on Sharing All Influenza Data (GISAID) database is higher, requiring that greater than 90% of the genome is covered by confident calls and that there is no evidence of contamination.

### Clustering and phylogenetic analysis

Lineages (Rambaut *et al*., 2020) assigned to each consensus genome were determined using Pangolin (https://github.com/cov-lineages/pangolin) from the Rambaut group on MRC CLIMB (Connor *et al*., 2016) using pangoLEARN training data set from 2020.08.29, with uncertain classifications (low support or discordance from tree) curated by hand. Only samples that passed quality control were considered for genomic analysis. Global sequences (Supplementary Table 2) that were potentially related to those from Zimbabwe were included in the phylogenetic analysis, leading to 6467 genomes being included using the software peroba (https://github.com/quadram-institute-bioscience/peroba). All sequences were aligned against the reference Wuhan Hu-1 using MAFFT (Katoh and Standley, 2013), and the maximum likelihood tree was estimated using fasttree and refined with iqtree2 under the HKY+G model (Price *et al*., 2010; Minh *et al*., 2019). Using PastML (Ishikawa *et al*., 2019) for ancestral state reconstruction, local outbreaks were found and redundant global sequences were removed for visualisation purposes.

## Results

### Epidemiology of SARS-CoV-2 infections in Zimbabwe from March to October 2020

Epidemiological analysis was based on 8,099 positive cases identified from the 92,299 national samples made available between the 20^th^ March (first case) and 16^th^ October 2020. Bulawayo had the highest infection rate at 221 per 100,000 population, followed by Harare at 144 per 100,000 population and then Matabeleland South at 106 cases per 100,000 population. Manicaland, Harare and Bulawayo provinces had the highest case fatality rates (CFR): 4.4%, 3.7% and 3.0% of the population, respectively (Table 1). Matabeleland South had the lowest CFR: 0.9%. The country’s overall CFR rate was 3% with almost half of the deaths (47%) happened in the community rather than in hospitals.

The majority of cases recorded prior to 20^th^ of July were travel associated. The number of cases with no recent travel reported remained below 50 during this period of time. People travelling from South Africa accounted for the greatest proportion (83.4%) of travel-associated cases (Fig. 1). From the time the first case was recorded to about 120 days later, there were more travel-associated cases than locally-transmitted cases, and the majority of travel-related cases were asymptomatic. During July the number of cases with no recent travel reported increased rapidly, confirming the start of community transmission. An increase in overall number of daily cases was observed from day 60 after the first case and peaked during the first half of August. Thereafter the number of cases decreased to the middle of October when approximately 20 cases per day were identified.

**Figure 1.**
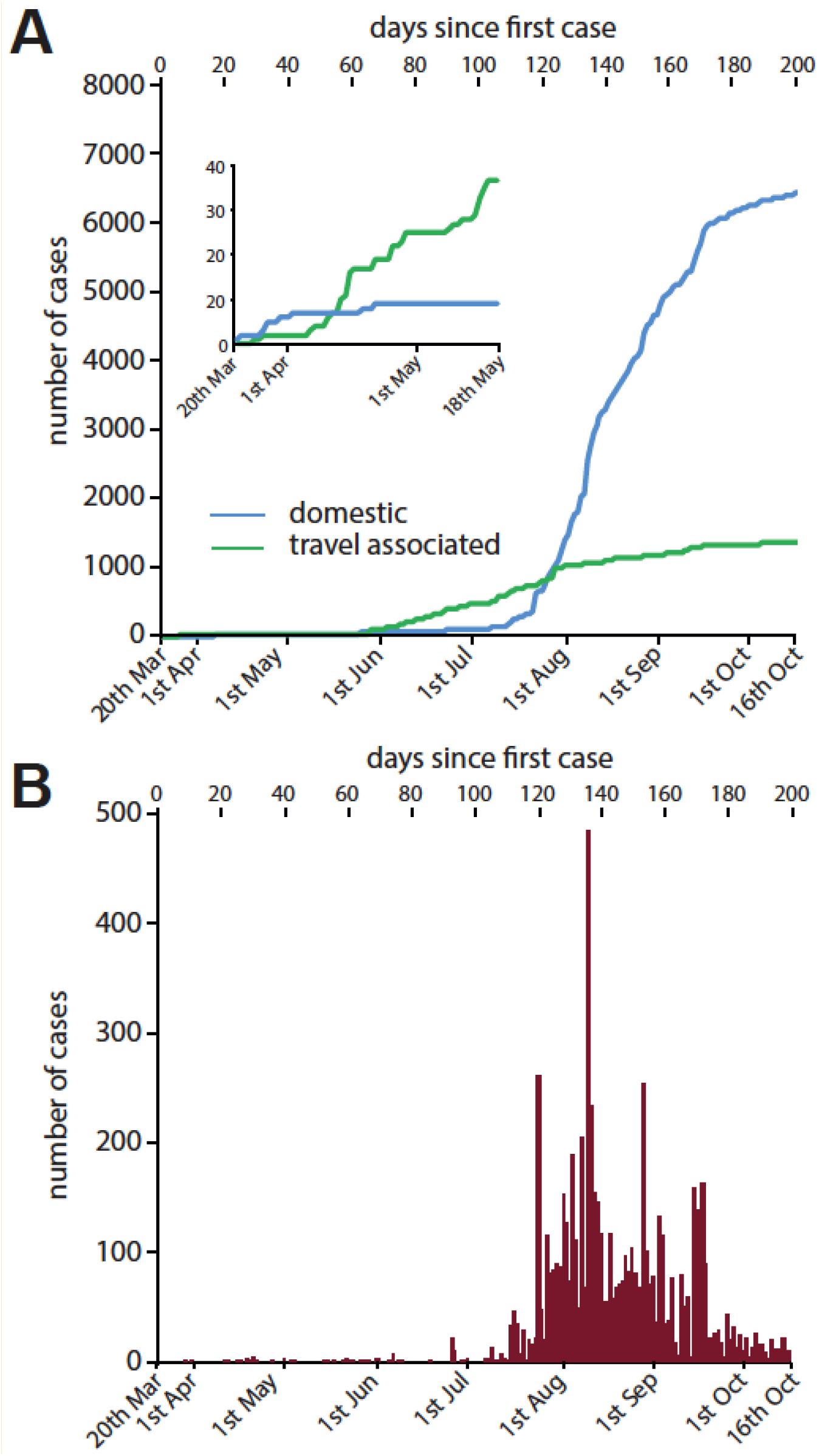
Epidemic curve of SARS-CoV-2 confirmed cases as of 16 October 2020. (**A**) Cumulative number of positive tests for SARS-CoV-2 in individuals reporting recent travel (probable imported cases, green line) or no recent travel (domestic cases, blue line). (**B**) Number of positive tests for SARS-CoV-2 by day (red bar).

Of those infected, 4455 (55%) were men and 3644 (45%) were women. Health workers constituted about 8% of the total number of reported cases. The most affected age group was the 31-40 years group (Fig. 2). The proportion of cases recorded in all age group categories were slightly greater amongst men than women except in the 91-100 years age group (Fig. 2). The greatest skew towards men was in the age range 41-50 while in females it was amongst the 91-100 age group. Overall, most deaths occurred amongst men (59%) and the largest number of deaths were in the 41-50 years age group. The CFR increased from age-group 41-50 to >90 years. The under-1-year CFR was 5% (Fig. 2).

**Figure 2.**
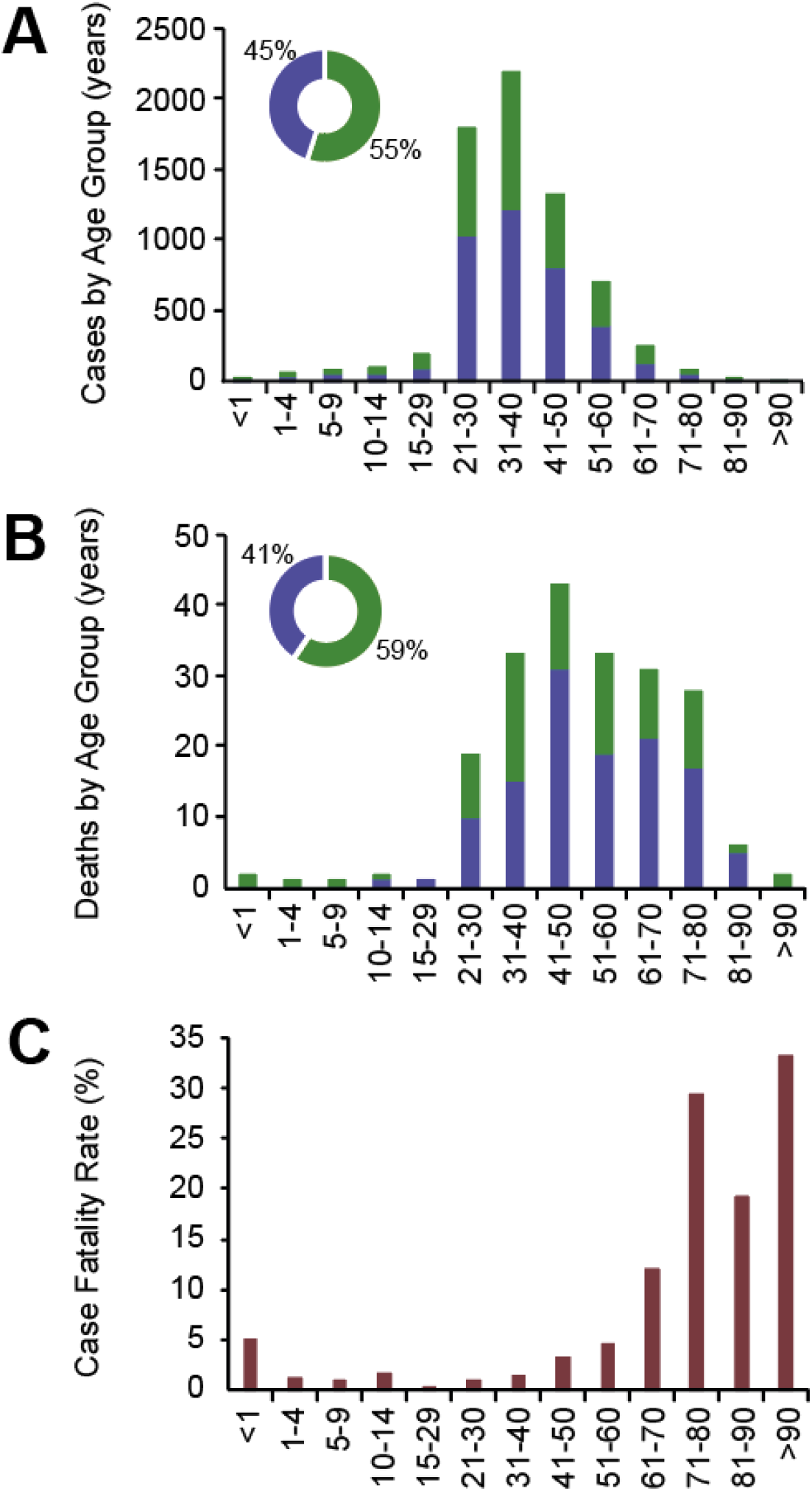
Relationship between age, gender and infection with SARS-Cov-2. (**A**) number of positive cases in each age group. (**B**) number of deaths in each age-group. (**C**) overall case fatality rate (CFR) in each age group. Cases for female (Blue bars) and male (green bar) are indicated. The small infographic in **A** and **B** represents the overall percentage of each gender infected (blue = female, green = male).

### At least 25 independent introductions of SARS-CoV-2 into Zimbabwe was associated with eight global lineages

The whole genome sequence of 100 SARS-CoV-2 samples from the first 3 months of the epidemic in Zimbabwe were analysed to investigate their phylogenetic relationship and infer spread. To place the genomes from Zimbabwe SARS-CoV-2 samples into the context of closely-related genomes from global samples a maximum likelihood phylogenetic tree of 6467 genomes and subsamples was constructed (Figure 3). The 100 genomes from Zimbabwe could be placed within eight previously defined high-order global sub-lineages (Table 2). Four samples (4%), were from high-order lineage A, with the remaining genomes (n=96, 96%) from lineage B. The most common sub-lineage was B.1.1 (51% of samples), followed by lineage B.1 (25% of samples) (Table 2).

**Figure 3.**
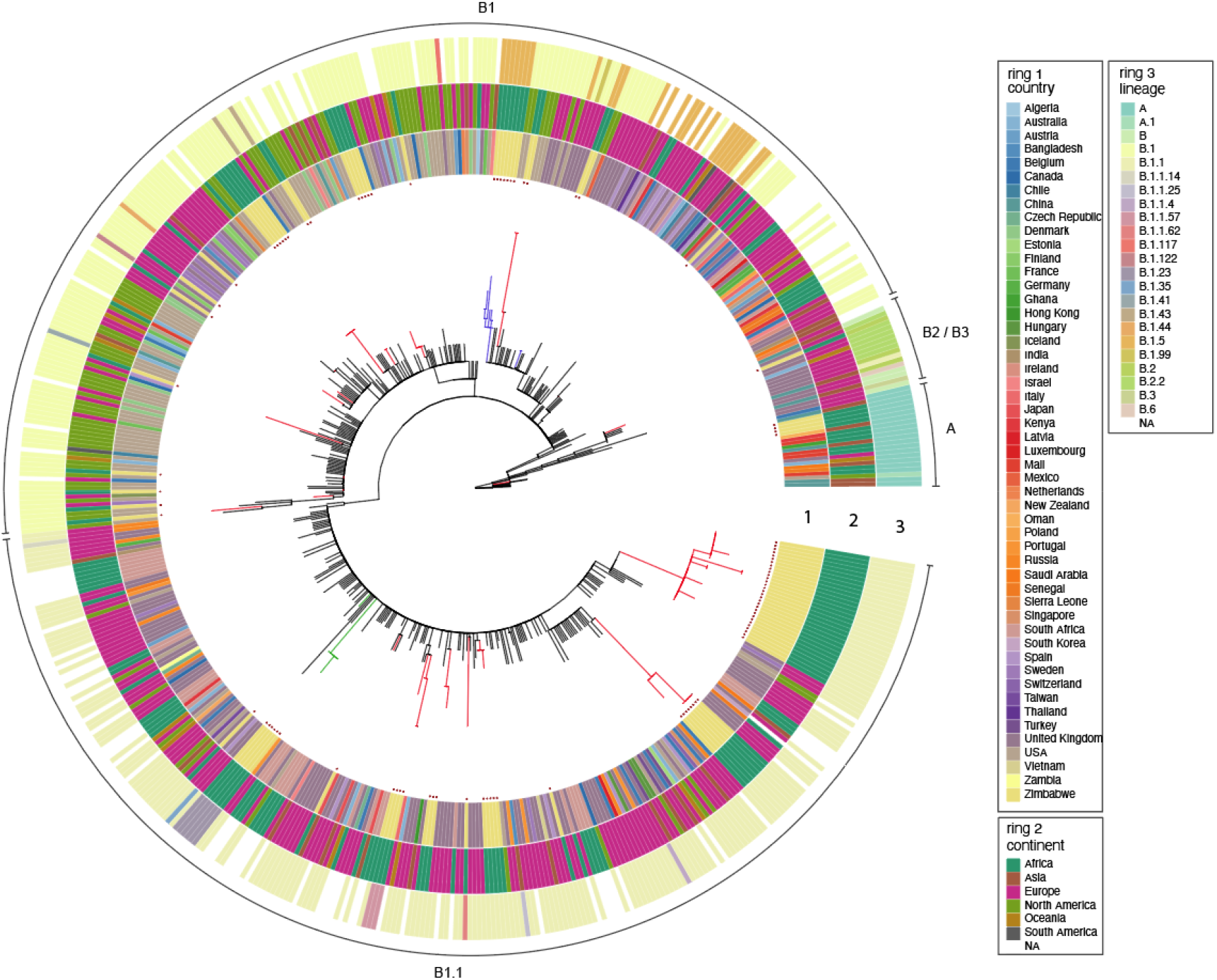
Phylogenetic relationship of SARS-CoV-2 genomes from Zimbabwe in context with closely related genomes from global cases. Maximum Likelihood phylogenetic tree of SARS-CoV-2 genomes from Zimbabwe in the context of 465 of the most closely-related genomes from global samples. Lineages connecting Zimbabwe clusters (red) and global lineages (black) are indicated. Country (inner circle) and continent (middle circle) of sampling, and global lineage designation (outer circle) are colour coded as indicated in the key (insert).

A maximum likelihood tree based on variation in the nucleotide sequence revealed a population structure for Zimbabwean cases of COVID-19 that grouped into 12 clusters each comprised of multiple genomes that emerged from nodes distributed throughout the tree; 15 genomes were present as a separate lineage (Figure 3). If each of these clusters represented separate introductions then we can infer that 27 was the lower limit for the number of independent introductions evident and for which there was good supporting evidence. However, in the case of four clusters or single genome lineages, the tree topology suggested that the genomes arose by divergence from just two introductions into Zimbabwe. In each case, two lineages shared a common ancestor with no intermediary nodes formed with genomes from other countries (blue and green lineages in Figure 3). By this more conservative estimation, the minimum number of introductions into Zimbabwe that our data support was 25. The 12 clusters were each comprised between 2 and 30 genomes with 0-3 SNPs between members of the cluster. Seven clusters contained more than one genome that had an identical genome sequence to at least one genome in the same cluster.

Two large clusters within lineage B.1.1 were present on extended branches of the phylogenetic tree composed of genomes that were distinct from all other to any genome present in the GISAID database by 4-5 SNPs. These introductions may have been from geographical locations where sequence of SARS-CoV-2 was not reported or the result of sequence divergence during transmission within Zimbabwe from an earlier case within Zimbabwe not included in our analysis.

### The D614G genotype was the most frequent variant in early spread events in Zimbabwe

There has been considerable interest in the emergence of a variant lineage of SARS-CoV-2 with a non-synonymous mutation resulting in substitution of an aspartate with a glycine residue at position 614 (D614G) of the spike (S) protein; this variant is linked to increased transmissibility in the upper respiratory tract and increased fitness (Plante *et al*., 2020; Volz *et al*., 2020). We therefore determined the distribution and frequency of the D614G variant in SARS-Cov-2 genomes from Zimbabwe. A total of 95 samples out of 100 contained the D614G mutation. Four of the five samples with the ancestral 614D form were from a single cluster of identical genomes from lineage A which was associated with travel from Dubai; the other was from lineage B.2 and associated with travel from USA. All five were from cases presenting in April or May 2020.

### Genomic epidemiology identifies intercontinental transmission and local transmission in the first 120 days of the epidemic

National surveillance in Zimbabwe identified the first three cases of COVID-19 in the second half of March 2020 in three individuals with a recent history of travel from outside of Zimbabwe. These patients had arrived from the United Kingdom (UK), United States (US) and Dubai and corresponded to samples ZW25, ZW29 and ZW70, respectively (Figure 4). Samples ZW25 and ZW29 were indistinguishable from one another suggesting a recent common source or direct transmission. However, this genome sequence was extremely common in the GISAID database (Shu and Mccauley, 2017) where identical genomes have been reported from multiple countries, although predominantly Europe, consistent with rapid transmission of this clone internationally (Figure 4).

**Figure 4.**
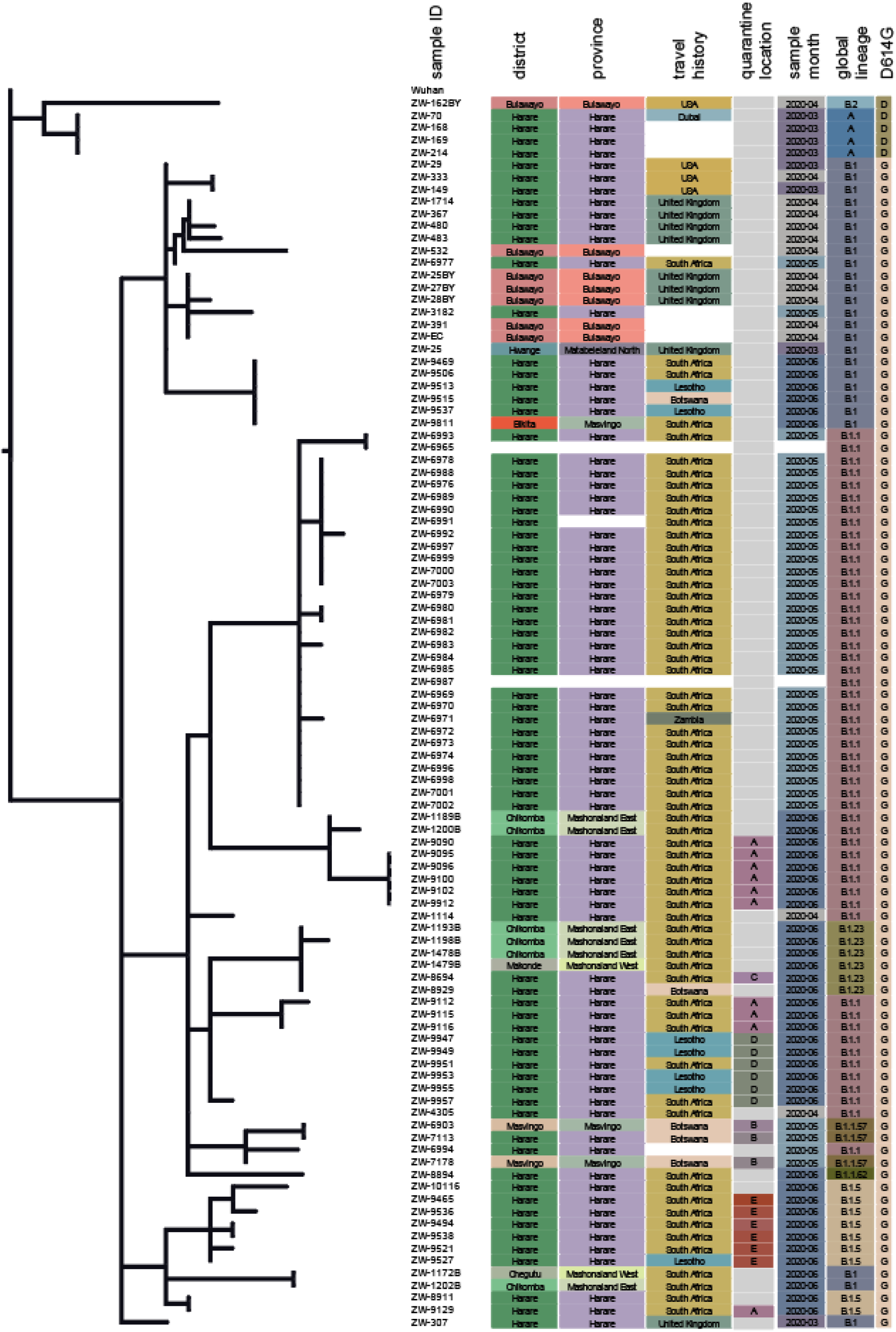
Phylogenetic relationship amongst 100 SARS-Cov-2 samples from Zimbabwe. A maximum likelihood tree based on SNPs in the genome of 100 SARS-Cov-2 genomes from Zimbabwe from between March and June 2020. The tree is rooted with the original SARS-Cov-2 genome from Wuhan. Sample Identifier, district and province location from which the sample was collected, country of travel if applicable or domestic case, quarantine facility identity (A-E) or no quarantine (white), month of isolation, SARS-Cov-2 global lineage and residue 614 of the spike ‘S’ protein are indicated in columns.

Genome ZW70 was from a traveller arriving from Dubai and was identical to three other genomes: ZW168, ZW169 and ZW214. Local epidemiological investigation indicated that these were infections resulting from cohabitation and may represent the earliest cases of local transmission. This interpretation was supported by the genome sequence data. Dubai airport is an important hub for flights from many locations in the world including Asia. Genomes ZW168, ZW169 and ZW214 all belonged to lineage A, which is most widely distributed in China.

An additional seven and three genomes were associated with arrivals from the UK and the US, respectively, in either late March or April. International travel was suspended following the arrival of these cases. Nine of these genomes belonged to the B1 lineage, while the last from a case associated with travel to the US belonged to the B2 lineage. Although in the same B1 lineage, eight of these travel-associated cases were on different sublineage and, with one exception, had no identical genomes from non-travel associated COVID-19 cases in our dataset. The one exception was sample ZW25BY that was from a case recorded in Bulawayo in April, which was identical to samples ZW-391 and ZW-EC, also from Bulawayo, with no travel-history recorded which, therefore, represents potential domestic transmission.

An increase in cases associated with Zimbabwean residents returning from neighbouring countries, especially South Africa, during May 2020 was reported by local surveillance. At this time SARS-CoV-2 was reported to be spreading rapidly in South Africa and therefore a two-week quarantine in designated accommodation was introduced. Although the majority of returnees were asymptomatic, they were tested and on multiple occasions were positive for SARS-CoV-2. A total of 26 samples from quarantined individuals housed in six of these accommodations were sequenced. In several quarantine accommodation settings, genomes from multiple samples from different individuals resident in the same quarantine location were identical or differed by only a single nucleotide in the genome. For example in quarantine location A, two clusters were identified that consisted of 10 and 14 samples (May 2020), respectively all of which were in the B1.1 lineage with genomes that were indistinguishable from one another; this was consistent with direct transmission. Since returnees from multiple locations were quarantined together, the most likely explanation is that transmission occurred in the quarantine accommodation rather than from a common outside source. Later spread events were primarily linked to returnees from South Africa and potential transmission in quarantine facilities.

## Discussion

Epidemiological analysis identified two phases of the SARS-CoV-2 epidemic in Zimbabwe. The vast majority of cases in the first 3 months were associated with international or regional travel. This was confirmed by the epidemiological data linking most confirmed cases with a history of having travelled outside of the country exacerbated by rapid influx of Zimbabwean citizens returning from other countries already experiencing the disease. A rapid increase in the number of cases was associated with probable domestic transmission after July 2020.

Zimbabwe is a landlocked country in Southern Africa between the Zambezi and Limpopo rivers and bordering South Africa, Botswana, Zambia and Mozambique. Travel history is essential for understanding the introduction of SARS-Cov-2 into a nation, and this was confirmed by the data from Zimbabwe. Initial cases recorded within the first 120 days of the outbreak were associated with travel from international locations already experiencing the disease. The majority of the initial COVID-19 cases in Zimbabwe (20^th^ March – 30^th^ June 2020) were Zimbabwean residents returning from neighbouring South Africa, Botswana and Mozambique. Most were associated with travel from South Africa related to informal trade between these two countries by cross-border traders, the majority of which are aged 25-45 years. The combination of informal cross-border traders and formally returning residents may have contributed to the subsequent increased local transmission in Zimbabwe. Other cases, however, were from Europe, Asia and USA, the epicentres of the SARS-Cov-2 epidemic. However, the rise is cases was slower than predicted and mirrored what has been seen in many African countries (Nkengasong and Mankoula, 2020). This slow rise in cases could be attributed to the measures the country instituted which included wearing of face masks, hand sanitization and lockdown. These measures have been shown to minimise and contain the outbreak in other countries, including France (Cauchemez *et al*., 2020).

The SARS-Cov-2 pandemic is an emerging infectious disease with no known public health interventions except the measures implemented in China, Europe and elsewhere. Zimbabwe’s response was shaped by experiences from other countries, where the epidemic was described first (La Regina *et al*., 2020; Shekhar *et al*., 2020; Xu *et al*., 2020). For the first 120 days following the first case of Covid-19 in Zimbabwe the total number of detected cases remained below 50, suggesting that public health interventions were effective in limiting local transmission to a minimum. The observed delayed local transmission may be as a result of the surveillance systems employed. Limiting movement by traders and cargo to a small manageable number, implementation of preventative equipment (e.g. hand sanitization, face mask wearing) and response measures within the first 30 days after the first case could explain this phenomenon. Though measures to minimise transmission were in place, a lot of irregularities emerged such as residents crossing borders illegally and/or non-compliance of quarantine requirements and relaxation of lockdown measures which could have contributed to an increase in local transmission. This was also evidenced by having the greatest number of cases centred around Bulawayo, Gweru and Harare cities with close proximity to land borders or international airports and densely populations. These factors may explain the propagated picture of the COVID-19 epidemic curve in Zimbabwe. Nonetheless, current evidence suggests that quarantine of travel-associated cases and early suspension of tourism-associated travel may have been effective based on our genomic analysis of cases in the first 120 days following the first case.

The factors associated with contraction of COVID-19 in Zimbabwe were consistent with factors associated with contraction in high income countries. The overall CFR of SARS Cov-2 was 2.9%, which was comparable with CFRs reported elsewhere in Asia, Europe and USA (Niforatos *et al*., 2020; Pachetti *et al*., 2020; Samaddar *et al*., 2020). Compared with other countries, younger males aged 31-40 years, were infected with SARS-CoV-2 in Zimbabwe during the first 120 days of the epidemic. However, the older population above 60 years was more likely to die from COVID-19 disease compared with the younger age groups, this agrees with what have been observed in other regions (Brandén *et al*., 2020; Dhama *et al*., 2020). Outbreak descriptions from Europe, Asia and the United States of America, indicated that older members of the population with pre-existing medical conditions were at increased risk of both becoming infected with SARS-Cov-2 and of dying from infection (Sanyaolu *et al*., 2020; Schultze *et al*., 2020). In the United Kingdom (UK) a considerably larger proportion of cases from ages 10-50 were in females, with cases in males being greatest in 0-10 years and 60-80 age categories (Page *et al*., 2020). These observations may have been due to two demographic characteristics prevalent in Zimbabwe. First, the younger age-group, 31-50 years is the most mobile and some have migrated out of the country in search of employment (Potts, 2010; Crush *et al*., 2015). Secondly, the same age group are sexually active and contribute the highest prevalence’s of HIV/ AIDS infection in the Southern Africa region (Gouws *et al*., 2008).

Fourteen out of the 100 SARS-Cov-2 cases sequenced reported that they had travelled from outside the Southern Africa region. Of these cases two resulted from local transmission with a positive epidemiological link. Amongst the remaining twelve cases there was no evidence of onward transmission based on the SARS-CoV-2 genomes sampled and sequenced in the subsequent two months. The sequences of SARS-CoV-2 genomes associated with travellers returning from neighbouring Southern African countries were distinct from those associated with travellers from the US, UK and Dubai recorded earlier in the epidemic; most were from lineage B1.1, which was also the most common lineage amongst 2016 sequences from 18 African countries reported recently (www.afro.who.int). Interestingly, lineage B.1.1.1 which accounted for 16% of the samples reported in South Africa was not found in our analysis. This may be due to the fact that South Africa is a major airline hub in Southern Africa, connecting most of the countries to Europe, Asia and USA.

The D614G genotype was the most frequent variant in early spread events in Zimbabwe, consistent with the reported trend for increased frequency of this genotype in many locations globally, even when the ancestral genotype (614D) was previously established (Korber *et al*., 2020). There has been considerable interest in the emergence of a variant lineage of SARS-CoV-2 with a non-synonymous mutation resulting in substitution of 614G in which a glycine residue was replaced by an aspartate residue at position 614 (D614G) of the spike (S) protein. The S-protein is thought to be important for entry of the virus into host cells and D614G has been implicated in increased cell entry (Korber *et al*., 2020); the mechanism for this was via increasing the likelihood of binding the ACE2 receptor on the cell surface (Yurkovetskiy *et al*., 2020). Consistent with this idea, population genetic modelling studies indicated that the 614G variant increased in frequency in the UK relative to the 614D ancestral variant, suggesting a selective advantage for 614G (Volz *et al*., 2020). This protein is also the target in anti-viral (remdesivir and favipiravir) treatment trials for COVID-19 (Saxena, 2020). The mutation of SARS-CoV-2 is important because vaccine-escape strains may emerge quickly and there is also the potential for detection-failure if the target used in the RT-PCR tests for SARS-CoV-2 may accumulate mutations affecting the test. A total of 95 of the 100 SARS-CoV-2 genomes used in our analysis contained the D614G mutation. All five of the genomes with the ancestral 614D genotype were from samples from March or April 2020, with none observed in sequences from samples taken in May or June. The dominance of the 614G genotype in Zimbabwe likely reflects the relatively high frequency of the 614G genotype in countries from which the virus was introduced. However, since it has been suggested that the 614G genotype is present in greater numbers in the upper respiratory tract in infected people it may also have been more likely to spread once introduced into the Zimbabwean population.

A combination of local surveillance and whole genome sequencing revealed the early intercontinental and subsequent local international spread of the SARS-CoV-2 into Zimbabwe. The analysis provides insights into the impact of interventions on transmission and the basis for improved future responses to this and other pandemics. Our whole genome sequence analysis provides a baseline view of the SARS-CoV-2 introduced into Zimbabwe that will place later domestic spread of the virus within a Zimbabwean context. As the national response to the SARS-CoV-2 pandemic continues the capability to test, isolate and track the evolution of SARS-CoV-2, is crucial to assess ongoing mutation of the virus.

## Conclusion

Phylogenomic analysis highlights the dominance of regional migration over intercontinental travel in the spread of SARS-CoV-2 in Zimbabwe. The majority of SARS-CoV-2 genomes from lineages B.1 and B.1.1 were recovered from residents returning from South Africa. These lineages appear to be predominant in Africa and continuous efforts for widespread sequencing is important to provide insights about their origin, and the evolution of local/imported viral dissemination in Zimbabwe. As the epidemic unfolded most of the more than 8,000 cases were due to local transmission. Risk factors for local transmission have not been described. Continuous surveillance using whole-genome sequencing is required to estimate the presence of mutations that may be increasing transmissibility. In addition to describing the immunological response to SARS-CoV-2 infection, data from SARS-CoV-2 genomic surveillance may contribute to the development of vaccines against this novel virus. Zimbabwe will need to strengthen health systems so that public health interventions that are effective against SARS-CoV-2 transmission elsewhere in the world, are also effective in Zimbabwe.

### Limitations

Samples used for whole genome sequencing where sampled from the first 120 days of the epidemic. Potential subsequent circulation of descendants of these early intercontinental introductions after around 120 days once transmission was increasing exponentially, remains to be determined.

## Data Availability

The data that support the findings of this study are openly available

## Ethical approval

Ethical approval for the study was exempted by the Medical Research Council of Zimbabwe as this study was in response to a public health emergency.

## Acknowledgements

The authors thank all local clinical and laboratory staff for their contribution and dedication to the SARS-CoV-2 Surveillance Network. Special thanks go to Elizabeth Tabitha Abbew, for all her expert advice and proofreading of the article. The authors acknowledge the Epidemiology and Diseases Control Department of the Ministry of Health and Child Care who provided full access to all the COVID-19 datasets exploited in this study. We thank members of the COVID-19 Genomics Consortium UK for their contributions to generating some of the data used in this study. We thank the laboratories who submitted their data to GISAID, full details are listed in the Supplementary Material.

## Author contributions

**Leadership and supervision:** AJP, RAK, JOG, SMZ

**Funding acquisition:** AJP, RAK, JOG

**Metadata curation:** TM, GT SMZ, GM, MGM

**Project administration:** TM, RAK, AJP, GM

**Samples and logistics:** TM, GT, AVG, TT, MGM, TA, HG, SARS-CoV-2 Research Group, Zimbabwe

**Sequencing and bioinformatics analysis:** DJB, AJP, LOM

**Data analysis**: TM, TT, LM, AP, RAK, IP, PM, AG, SD, IC, JJ, RM, KM, IM, DM, MH

**Paper writing**: TM, RAK, LM, JC, BVC, FTK, FTT, JC, MGM, GM, SMZ

**Appendix 1** (The COVID-19 Genomics UK (COG-UK) Consortium)

**Funding acquisition, leadership, supervision, metadata curation, project administration, samples, logistics, Sequencing, analysis, and Software and analysis tools:**

Thomas R Connor ^33, 34^, and Nicholas J Loman ^15^.

**Leadership, supervision, sequencing, analysis, funding acquisition, metadata curation, project administration, samples, logistics, and visualisation:**

Samuel C Robson ^68^.

**Leadership, supervision, project administration, visualisation, samples, logistics, metadata curation and software and analysis tools:**

Tanya Golubchik ^27^.

**Leadership, supervision, metadata curation, project administration, samples, logistics sequencing and analysis:**

M. Estee Torok ^8, 10^.

**Project administration, metadata curation, samples, logistics, sequencing, analysis, and software and analysis tools:**

William L Hamilton ^8, 10^.

**Leadership, supervision, samples logistics, project administration, funding acquisition sequencing and analysis:**

David Bonsall ^27^.

**Leadership and supervision, sequencing, analysis, funding acquisition, visualisation and software and analysis tools:**

Ali R Awan ^74^.

**Leadership and supervision, funding acquisition, sequencing, analysis, metadata curation, samples and logistics:**

Sally Corden^33^.

**Leadership supervision, sequencing analysis, samples, logistics, and metadata curation:** Ian Goodfellow ^11^.

**Leadership, supervision, sequencing, analysis, samples, logistics, and Project administration:**

Darren L Smith ^60, 61^.

**Project administration, metadata curation, samples, logistics, sequencing and analysis:**

Martin D Curran ^14^, and Surendra Parmar ^14^.

**Samples, logistics, metadata curation, project administration sequencing and analysis:**

James G Shepherd ^21^.

**Sequencing, analysis, project administration, metadata curation and software and analysis tools:**

Matthew D Parker ^38^ and Dinesh Aggarwal ^1, 2, 3^.

**Leadership, supervision, funding acquisition, samples, logistics, and metadata curation:**

Catherine Moore ^33^.

**Leadership, supervision, metadata curation, samples, logistics, sequencing and analysis:**

Derek J Fairley^6, 88^, Matthew W Loose ^54^, and Joanne Watkins ^33^.

**Metadata curation, sequencing, analysis, leadership, supervision and software and analysis tools:**

Matthew Bull ^33^, and Sam Nicholls ^15^.

**Leadership, supervision, visualisation, sequencing, analysis and software and analysis tools:**David M Aanensen ^1, 30^.

**Sequencing, analysis, samples, logistics, metadata curation, and visualisation:**

Sharon Glaysher ^70^.

**Metadata curation, sequencing, analysis, visualisation, software and analysis tools:**

Matthew Bashton ^60^, and Nicole Pacchiarini ^33^.

**Sequencing, analysis, visualisation, metadata curation, and software and analysis tools**:

Anthony P Underwood ^1, 30^.

**Funding acquisition, leadership, supervision and project administration:**

Thushan I de Silva ^38^, and Dennis Wang ^38^.

**Project administration, samples, logistics, leadership and supervision**:

Monique Andersson^28^, Anoop J Chauhan ^70^, Mariateresa de Cesare ^26^, Catherine Ludden

^1,3^, and Tabitha W Mahungu ^91^.

**Sequencing, analysis, project administration and metadata curation:**

Rebecca Dewar ^20^, and Martin P McHugh ^20^.

**Samples, logistics, metadata curation and project administration:**

Natasha G Jesudason ^21^, Kathy K Li MBBCh ^21^, Rajiv N Shah ^21^, and Yusri Taha ^66^.

**Leadership, supervision, funding acquisition and metadata curation:**

Kate E Templeton ^20^.

**Leadership, supervision, funding acquisition, sequencing and analysis:**

Simon Cottrell ^33^, Justin O’Grady ^51^, Andrew Rambaut ^19^, and Colin P Smith^93^.

**Leadership, supervision, metadata curation**, **sequencing and analysis:**

Matthew T.G. Holden ^87^, and Emma C Thomson ^21^.

**Leadership, supervision, samples, logistics and metadata curation**: Samuel Moses ^81, 82^.

**Sequencing, analysis, leadership, supervision, samples and logistics:**

Meera Chand ^7^, Chrystala Constantinidou ^71^, Alistair C Darby ^46^, Julian A Hiscox ^46^, Steve Paterson ^46^, and Meera Unnikrishnan ^71^.

**Sequencing, analysis, leadership and supervision and software and analysis tools:**

Andrew J Page ^51^, and Erik M Volz ^96^.

**Samples, logistics, sequencing, analysis and metadata curation:**

Charlotte J Houldcroft ^8^, Aminu S Jahun ^11^, James P McKenna ^88^, Luke W Meredith ^11^, Andrew Nelson ^61^, Sarojini Pandey ^72^, and Gregory R Young ^60^.

**Sequencing, analysis, metadata curation, and software and analysis tools:**

Anna Price ^34^, Sara Rey ^33^, Sunando Roy ^41^, Ben Temperton^49^, and Matthew Wyles ^38^.

**Sequencing, analysis, metadata curation and visualisation:**

Stefan Rooke^19^, and Sharif Shaaban ^87^.

**Visualisation, sequencing, analysis and software and analysis tools:**

Helen Adams ^35^, Yann Bourgeois ^69^, Katie F Loveson ^68^,Áine O’Toole ^19^, and Richard Stark ^71^.

**Project administration, leadership and supervision:**

Ewan M Harrison ^1, 3^, David Heyburn ^33^, and Sharon J Peacock ^2, 3^

**Project administration and funding acquisition:**

David Buck ^26^, and Michaela John^36^

**Sequencing, analysis and project administration:**

Dorota Jamrozy ^1^, and Joshua Quick ^15^

**Samples, logistics, and project administration:**

Rahul Batra ^78^, Katherine L Bellis ^1, 3^, Beth Blane ^3^, Sophia T Girgis ^3^, Angie Green ^26^, Anita Justice ^28^, Mark Kristiansen ^41^, and Rachel J Williams ^41^.

**Project administration, software and analysis tools:**

Radoslaw Poplawski^15^.

**Project administration and visualisation:**

Garry P Scarlett ^69^.

**Leadership, supervision, and funding acquisition:**

John A Todd ^26^, Christophe Fraser ^27^, Judith Breuer ^40,41^, Sergi Castellano ^41^, Stephen L Michell ^49^, Dimitris Gramatopoulos ^73^, and Jonathan Edgeworth ^78^.

**Leadership, supervision and metadata curation:**

Gemma L Kay ^51^.

**Leadership, supervision, sequencing and analysis:**

Ana da Silva Filipe ^21^, Aaron R Jeffries ^49^, Sascha Ott ^71^, Oliver Pybus ^24^, David L Robertson ^21^, David A Simpson ^6^, and Chris Williams ^33^.

**Samples, logistics, leadership and supervision:**

Cressida Auckland ^50^, John Boyes ^83^, Samir Dervisevic ^52^, Sian Ellard ^49, 50^, Sonia Goncalves^1^, Emma J Meader ^51^, Peter Muir ^2^, Husam Osman ^95^, Reenesh Prakash ^52^, Venkat Sivaprakasam ^18^, and Ian B Vipond ^2^.

**Leadership, supervision and visualisation**

Jane AH Masoli ^49, 50^.

**Sequencing, analysis and metadata curation**

Nabil-Fareed Alikhan ^51^, Matthew Carlile ^54^, Noel Craine ^33^, Sam T Haldenby ^46^, Nadine Holmes ^54^, Ronan A Lyons ^37^, Christopher Moore ^54^, Malorie Perry ^33^, Ben Warne ^80^, and Thomas Williams ^19^.

**Samples, logistics and metadata curation:**

Lisa Berry ^72^, Andrew Bosworth ^95^, Julianne Rose Brown ^40^, Sharon Campbell ^67^, Anna Casey ^17^,Gemma Clark ^56^, Jennifer Collins ^66^, Alison Cox ^43, 44^, Thomas Davis ^84^, Gary Eltringham ^66^, Cariad Evans ^38, 39^, Clive Graham ^64^, Fenella Halstead ^18^, Kathryn Ann Harris ^40^, Christopher Holmes ^58^, Stephanie Hutchings ^2^, Miren Iturriza-Gomara ^46^, Kate Johnson ^38, 39^, Katie Jones ^72^, Alexander J Keeley ^38^, Bridget A Knight ^49, 50^, Cherian Koshy^90^, Steven Liggett ^63^, Hannah Lowe ^81^, Anita O Lucaci ^46^, Jessica Lynch ^25, 29^, Patrick C McClure ^55^, Nathan Moore ^31^, Matilde Mori ^25, 29, 32^, David G Partridge ^38, 39^, Pinglawathee Madona ^43, 44^, Hannah M Pymont ^2^, Paul Anthony Randell ^43, 44^, Mohammad Raza ^38, 39^, Felicity Ryan ^81^, Robert Shaw ^28^, Tim J Sloan ^57^, and Emma Swindells ^65^.

**Sequencing, analysis, Samples and logistics:**

Alexander Adams ^33^, Hibo Asad ^33^, Alec Birchley ^33^, Tony Thomas Brooks ^41^, Giselda Bucca ^93^, Ethan Butcher ^70^, Sarah L Caddy ^13^, Laura G Caller ^2, 3, 12^, Yasmin Chaudhry ^11^, Jason Coombes ^33^, Michelle Cronin ^33^, Patricia L Dyal ^41^, Johnathan M Evans ^33^, Laia Fina ^33^, Bree Gatica-Wilcox ^33^, Iliana Georgana ^11^, Lauren Gilbert ^33^, Lee Graham ^33^, Danielle C Groves ^38^, Grant Hall ^11^, Ember Hilvers ^33^, Myra Hosmillo ^11^, Hannah Jones ^33^, Sophie Jones ^33^, Fahad A Khokhar ^13^, Sara Kumziene-Summerhayes ^33^, George MacIntyre-Cockett ^26^, Rocio T Martinez Nunez ^94^, Caoimhe McKerr ^33^, Claire McMurray ^15^, Richard Myers ^7^, Yasmin Nicole Panchbhaya ^41^, Malte L Pinckert ^11^, Amy Plimmer ^33^, Joanne Stockton ^15^, Sarah Taylor ^33^, Alicia Thornton ^7^, Amy Trebes ^26^, Alexander J Trotter ^51^, Helena Jane Tutill ^41^, Charlotte A Williams ^41^, Anna Yakovleva ^11^ and Wen C Yew ^62^.

**Sequencing, analysis and software and analysis tools:**

Mohammad T Alam ^71^, Laura Baxter ^71^, Olivia Boyd ^96^, Fabricia F. Nascimento ^96^, Timothy M Freeman ^38^, Lily Geidelberg ^96^, Joseph Hughes ^21^, David Jorgensen ^96^, Benjamin B Lindsey ^38^, Richard J Orton ^21^, Manon Ragonnet-Cronin ^96^ Joel Southgate ^33, 34^, and Sreenu Vattipally ^21^.

**Samples, logistics and software and analysis tools:**

Igor Starinskij ^23^.

**Visualisation and software and analysis tools:**

Joshua B Singer ^21^, Khalil Abudahab ^1, 30^, Leonardo de Oliveira Martins ^51^, Thanh Le-Viet ^51^,Mirko Menegazzo ^30^, Ben EW Taylor ^1, 30^, and Corin A Yeats ^30^.

**Project Administration:**

Sophie Palmer ^3^, Carol M Churcher ^3^, Alisha Davies ^33^, Elen De Lacy ^33^, Fatima Downing ^33^, Sue Edwards ^33^, Nikki Smith ^38^, Francesc Coll ^97^, Nazreen F Hadjirin ^3^ and Frances Bolt ^44, 45^.

**Leadership and supervision:**

Alex Alderton^1^, Matt Berriman^1^, Ian G Charles ^51^, Nicholas Cortes ^31^, Tanya Curran ^88^, John Danesh^1^, Sahar Eldirdiri ^84^, Ngozi Elumogo ^52^, Andrew Hattersley ^49, 50^, Alison Holmes ^44, 45^, Robin Howe ^33^, Rachel Jones ^33^, Anita Kenyon ^84^, Robert A Kingsley ^51^, Dominic Kwiatkowski ^1, 9^, Cordelia Langford^1^, Jenifer Mason^48^, Alison E Mather ^51^, Lizzie Meadows ^51^, Sian Morgan ^36^, James Price ^44, 45^, Trevor I Robinson ^48^, Giri Shankar ^33^, John Wain ^51^, and Mark A Webber ^51^.

**Metadata curation:**

Declan T Bradley ^5, 6^, Michael R Chapman ^1, 3, 4^, Derrick Crooke ^28^, David Eyre ^28^, Martyn Guest ^34^, Huw Gulliver ^34^, Sarah Hoosdally ^28^, Christine Kitchen ^34^, Ian Merrick ^34^, Siddharth Mookerjee ^44, 45^, Robert Munn ^34^, Timothy Peto ^28^, Will Potter ^52^, Dheeraj K Sethi ^52^, Wendy Smith ^56^, Luke B Snell ^75, 94^, Rachael Stanley ^52^, Claire Stuart ^52^ and Elizabeth Wastenge^20^.

**Sequencing and analysis:**

Erwan Acheson ^6^, Safiah Afifi ^36^, Elias Allara ^2, 3^, Roberto Amato ^1^, Adrienn Angyal ^38^, Elihu Aranday-Cortes ^21^, Cristina Ariani ^1^, Jordan Ashworth ^19^, Stephen Attwood ^24^, Alp Aydin ^51^, David J Baker ^51^, Carlos E Balcazar ^19^, Angela Beckett ^68^ Robert Beer ^36^, Gilberto Betancor ^76^, Emma Betteridge ^1^, David Bibby ^7^, Daniel Bradshaw^7^, Catherine Bresner ^34^, Hannah E Bridgewater ^71^, Alice Broos ^21^, Rebecca Brown ^38^, Paul E Brown ^71^, Kirstyn Brunker ^22^, Stephen N Carmichael ^21^, Jeffrey K. J. Cheng ^71^, Dr Rachel Colquhoun ^19^, Gavin Dabrera ^7^, Johnny Debebe ^54^, Eleanor Drury ^1^, Louis du Plessis ^24^, Richard Eccles ^46^, Nicholas Ellaby ^7^, Audrey Farbos ^49^, Ben Farr ^1^, Jacqueline Findlay ^41^, Chloe L Fisher ^74^, Leysa Marie Forrest ^41^, Sarah Francois ^24^, Lucy R. Frost ^71^, William Fuller^34^, Eileen Gallagher ^7^, Michael D Gallagher ^19^, Matthew Gemmell ^46^, Rachel AJ Gilroy ^51^, Scott Goodwin ^1^, Luke R Green ^38^, Richard Gregory ^46^, Natalie Groves ^7^, James W Harrison ^49^, Hassan Hartman ^7^, Andrew R Hesketh ^93^,Verity Hill ^19^, Jonathan Hubb ^7^, Margaret Hughes^46^, David K Jackson ^1^, Ben Jackson ^19^, Keith James ^1^, Natasha Johnson ^21^, Ian Johnston ^1^, Jon-Paul Keatley ^1^, Moritz Kraemer ^24^, Angie Lackenby ^7^, Mara Lawniczak ^1^, David Lee ^7^, Rich Livett ^1^, Stephanie Lo ^1^, Daniel Mair ^21^, Joshua Maksimovic ^36^, Nikos Manesis ^7^, Robin Manley ^49^, Carmen Manso ^7^, Angela Marchbank ^34^, Inigo Martincorena ^1^, Tamyo Mbisa ^7^, Kathryn McCluggage ^36^, JT McCrone ^19^, Shahjahan Miah ^7^, Michelle L Michelsen ^49^, Mari Morgan ^33^, Gaia Nebbia ^78^,Charlotte Nelson ^46^, Jenna Nichols ^21^, Paola Niola ^41^, Kyriaki Nomikou ^21^, Steve Palmer ^1^, Naomi Park ^1^, Yasmin A Parr ^1^, Paul J Parsons ^38^, Vineet Patel ^7^, Minal Patel ^1^, Clare Pearson ^2, 1^, Steven Platt ^7^, Christoph Puethe ^1^, Mike Quail ^1^,Jayna Raghwani ^24^, Lucille Rainbow ^46^, Shavanthi Rajatileka ^1^, Mary Ramsay ^7^, Paola C Resende Silva ^41, 42^, Steven Rudder 51, Chris Ruis ^3^, Christine M Sambles ^49^, Fei Sang ^54^, Ulf Schaefer^7^, Emily Scher ^19^, Carol Scott ^1^, Lesley Shirley ^1^, Adrian W Signell ^76^, John Sillitoe ^1^, Christen Smith ^1^, Dr Katherine L Smollett ^21^, Karla Spellman ^36^, Thomas D Stanton ^19^, David J Studholme ^49^, Grace Taylor-Joyce ^71^, Ana P Tedim ^51^, Thomas Thompson ^6^, Nicholas M Thomson ^51^, Scott Thurston^1^, Lily Tong ^21^, Gerry Tonkin-Hill ^1^, Rachel M Tucker ^38^, Edith E Vamos ^4^, Tetyana Vasylyeva^24^, Joanna Warwick-Dugdale ^49^, Danni Weldon ^1^, Mark Whitehead ^46^, David Williams ^7^, Kathleen A Williamson ^19^,Harry D Wilson ^76^,Trudy Workman ^34^, Muhammad Yasir^51^, Xiaoyu Yu ^19^, and Alex Zarebski ^24^.

**Samples and logistics:**

Evelien M Adriaenssens ^51^, Shazaad S Y Ahmad ^2, 47^, Adela Alcolea-Medina ^59, 77^, John Allan ^60^, Patawee Asamaphan ^21^, Laura Atkinson ^40^, Paul Baker ^63^, Jonathan Ball ^55^, Edward Barton^64^, Mathew A Beale^1^, Charlotte Beaver^1^, Andrew Beggs ^16^, Andrew Bell ^51^, Duncan J Berger ^1^, Louise Berry. ^56^, Claire M Bewshea ^49^, Kelly Bicknell ^70^, Paul Bird ^58^, Chloe Bishop ^7^, Tim Boswell ^56^, Cassie Breen ^48^, Sarah K Buddenborg^1^, Shirelle Burton-Fanning ^66^, Vicki Chalker ^7^, Joseph G Chappell ^55^, Themoula Charalampous ^78, 94^, Claire Cormie^3^, Nick Cortes^29, 25^, Lindsay J Coupland ^52^, Angela Cowell ^48^, Rose K Davidson ^53^, Joana Dias ^3^, Maria Diaz ^51^, Thomas Dibling^1^, Matthew J Dorman^1^, Nichola Duckworth^57^, Scott Elliott^70^, Sarah Essex^63^, Karlie Fallon ^58^, Theresa Feltwell ^8^, Vicki M Fleming ^56^, Sally Forrest ^3^, Luke Foulser^1^, Maria V Garcia-Casado^1^, Artemis Gavriil ^41^, Ryan P George ^47^, Laura Gifford ^33^, Harmeet K Gill ^3^, Jane Greenaway ^65^, Luke Griffith^53^, Ana Victoria Gutierrez^51^, Antony D Hale ^85^, Tanzina Haque ^91^, Katherine L Harper ^85^, Ian Harrison ^7^, Judith Heaney ^89^, Thomas Helmer ^58^, Ellen E Higginson^3^, Richard Hopes ^2^, Hannah C Howson-Wells ^56^, Adam D Hunter ^1^, Robert Impey ^70^, Dianne Irish-Tavares ^91^, David A Jackson^1^, Kathryn A Jackson ^46^, Amelia Joseph ^56^, Leanne Kane ^1^, Sally Kay ^1^, Leanne M Kermack ^3^, Manjinder Khakh ^56^, Stephen P Kidd ^29, 25, 31^, Anastasia Kolyva ^51^, Jack CD Lee ^40^, Laura Letchford ^1^, Nick Levene ^79^, Lisa J Levett ^89^, Michelle M Lister ^56^, Allyson Lloyd ^70^, Joshua Loh ^60^, Louissa R Macfarlane-Smith ^85^, Nicholas W Machin ^2, 47^, Mailis Maes ^3^, Samantha McGuigan ^1^, Liz McMinn ^1^, Lamia Mestek-Boukhibar ^41^, Zoltan Molnar ^6^, Lynn Monaghan ^79^, Catrin Moore ^27^, Plamena Naydenova ^3^, Alexandra S Neaverson ^1^, Rachel Nelson ^1^, Marc O Niebel ^21^, Elaine O’Toole^48^, Debra Padgett ^64^, Gaurang Patel ^1^, Brendan AI Payne ^66^, Liam Prestwood ^1^, Veena Raviprakash ^67^, Nicola Reynolds^86^, Alex Richter ^16^, Esther Robinson ^95^, Hazel A Rogers^1^, Aileen Rowan ^96^, Garren Scott ^64^, Divya Shah ^40^, Nicola Sheriff ^67^, Graciela Sluga, Emily Souster^1^, Michael Spencer-Chapman^1^, Sushmita Sridhar ^1, 3^, Tracey Swingler ^53^, Julian Tang^58^, Graham P Taylor^96^, Theocharis Tsoleridis ^55^, Lance Turtle^46^, Sarah Walsh ^57^, Michelle Wantoch ^86^, Joanne Watts ^48^, Sheila Waugh ^66^, Sam Weeks^41^, Rebecca Williams^31^, Iona Willingham^56^, Emma L Wise ^25, 29, 31^, Victoria Wright ^54^, Sarah Wyllie ^70^, and Jamie Young ^3^.

**Software and analysis tools**

Amy Gaskin^33^, Will Rowe ^15^, and Igor Siveroni ^96^.

**Visualisation:**

Robert Johnson ^96^.

**Affiliations:**

**1** Wellcome Sanger Institute, **2** Public Health England, **3** University of Cambridge, **4** Health Data Research UK, Cambridge, **5** Public Health Agency, Northern Ireland, **6** Queen’s University Belfast **7** Public Health England Colindale, **8** Department of Medicine, University of Cambridge, **9** University of Oxford, **10** Departments of Infectious Diseases and Microbiology, Cambridge University Hospitals NHS Foundation Trust; Cambridge, UK, **11** Division of Virology, Department of Pathology, University of Cambridge, **12** The Francis Crick Institute, **13** Cambridge Institute for Therapeutic Immunology and Infectious Disease, Department of Medicine, **14** Public Health England, Clinical Microbiology and Public Health Laboratory, Cambridge, UK, **15** Institute of Microbiology and Infection, University of Birmingham, **16** University of Birmingham, **17** Queen Elizabeth Hospital, **18** Heartlands Hospital, **19** University of Edinburgh, **20** NHS Lothian, **21** MRC-University of Glasgow Centre for Virus Research, **22** Institute of Biodiversity, Animal Health & Comparative Medicine, University of Glasgow, **23** West of Scotland Specialist Virology Centre, **24** Dept Zoology, University of Oxford, **25** University of Surrey, **26** Wellcome Centre for Human Genetics, Nuffield Department of Medicine, University of Oxford, **27** Big Data Institute, Nuffield Department of Medicine, University of Oxford, **28** Oxford University Hospitals NHS Foundation Trust, **29** Basingstoke Hospital, **30** Centre for Genomic Pathogen Surveillance, University of Oxford, **31** Hampshire Hospitals NHS Foundation Trust, **32** University of Southampton, **33** Public Health Wales NHS Trust, **34** Cardiff University, **35** Betsi Cadwaladr University Health Board, **36** Cardiff and Vale University Health Board, **37** Swansea University, **38** University of Sheffield, **39** Sheffield Teaching Hospitals, **40** Great Ormond Street NHS Foundation Trust, **41** University College London, **42** Oswaldo Cruz Institute, Rio de Janeiro **43** North West London Pathology, **44** Imperial College Healthcare NHS Trust, **45** NIHR Health Protection Research Unit in HCAI and AMR, Imperial College London, **46** University of Liverpool, **47** Manchester University NHS Foundation Trust, **48** Liverpool Clinical Laboratories, **49** University of Exeter, **50** Royal Devon and Exeter NHS Foundation Trust, **51** Quadram Institute Bioscience, University of East Anglia, **52** Norfolk and Norwich University Hospital, **53** University of East Anglia, **54** Deep Seq, School of Life Sciences, Queens Medical Centre, University of Nottingham, **55** Virology, School of Life Sciences, Queens Medical Centre, University of Nottingham, **56** Clinical Microbiology Department, Queens Medical Centre, **57** PathLinks, Northern Lincolnshire & Goole NHS Foundation Trust, **58** Clinical Microbiology, University Hospitals of Leicester NHS Trust, **59** Viapath, **60** Hub for Biotechnology in the Built Environment, Northumbria University, **61** NU-OMICS Northumbria University, **62** Northumbria University, **63** South Tees Hospitals NHS Foundation Trust, **64** North Cumbria Integrated Care NHS Foundation Trust, **65** North Tees and Hartlepool NHS Foundation Trust, **66** Newcastle Hospitals NHS Foundation Trust, **67** County Durham and Darlington NHS Foundation Trust, **68** Centre for Enzyme Innovation, University of Portsmouth, **69** School of Biological Sciences, University of Portsmouth, **70** Portsmouth Hospitals NHS Trust, **71** University of Warwick, **72** University Hospitals Coventry and Warwickshire, **73** Warwick Medical School and Institute of Precision Diagnostics, Pathology, UHCW NHS Trust, **74** Genomics Innovation Unit, Guy’s and St. Thomas’ NHS Foundation Trust, **75** Centre for Clinical Infection & Diagnostics Research, St. Thomas’ Hospital and Kings College London, **76** Department of Infectious Diseases, King’s College London, **77** Guy’s and St. Thomas’ Hospitals NHS Foundation Trust, **78** Centre for Clinical Infection and Diagnostics Research, Department of Infectious Diseases, Guy’s and St Thomas’ NHS Foundation Trust, **79** Princess Alexandra Hospital Microbiology Dept., **80** Cambridge University Hospitals NHS Foundation Trust, **81** East Kent Hospitals University NHS Foundation Trust, **82** University of Kent, **83** Gloucestershire Hospitals NHS Foundation Trust, **84** Department of Microbiology, Kettering General Hospital, **85** National Infection Service, PHE and Leeds Teaching Hospitals Trust, **86** Cambridge Stem Cell Institute, University of Cambridge, **87** Public Health Scotland, 88 Belfast Health & Social Care Trust, **89** Health Services Laboratories, **90** Barking, Havering and Redbridge University Hospitals NHS Trust, **91** Royal Free NHS Trust, **92** Maidstone and Tunbridge Wells NHS Trust, **93** University of Brighton, **94** Kings College London, **95** PHE Heartlands, **96** Imperial College London, **97** Department of Infection Biology, London School of Hygiene and Tropical Medicine.

**Appendix 2** (SARS-CoV-2 Research Group, Zimbabwe)

**Samples and logistics:**

Kenneth Maeka^1^, Peter Gumbo^1^, Amini David^1^, Tariro Madamombe^1^, Lucy Sisya^1^, Shukusho D Fungai^1^, Mercy Marumure^1^, Admire Bwgirire^1^, Florence Malunga^1^, Norah Sukutayi-Vere^1^, Takudzwa Kawome^1^, Stanford Mupandasekwa^1^,Tanaka Sakubani^1^, Exavier Mazarura^1^, Babra Murwira^2^, Monalisa Mutimutema^2^, Joshua Mbanga^3^, Zephania Dhlamini^3^, Aleck Maunganidze^3^, Anita Dube^3^, Beauty Makamure^4^, Justin Mayini^4^, Tendai Washaya^4^, Janice Martins^4^, Isabel Mashita^4^, Happiness Mahove^4^, Norah Karonga^4^, Forget Makoga^4^, Learnmore Chawira^5^, Emmaculate Govore^5^, Chipo Berejena^6^, Arnold Mukaratirwa^6^, Ruhanya Vurayayi^6^, John Macharaga^7^, Samson Chomunacho^7^, Simbarashe Mapurisa^7^, Albert Mudungwe^7^, Davison Makwarimba^7^, Raymond Ncube^7^, Beverley Chinembiri^7^, Godfrey Mhondiwa^7^, Nelia Kativu^7^, Rutendo Muchemwa^7^ and Everlyn Choto^7^.

**Paper writing:**

Ruhanya Vurayayi^6^.

**Affiliations:**

**1** National Microbiology Reference Laboratory, Ministry of Health and Child Care, Zimbabwe, P.O. Box ST 749, Southerton, Harare, Zimbabwe, **2** National TB Reference Laboratory, Bulawayo, **3** National University of Science and Technology, Bulawayo, **4** Biomedical Research and Training Institute, **5** Harare City Health Department, Beatrice Road Infectious Diseases Hospital, Harare, Zimbabwe, **6** National Virology Laboratory, Harare, Zimbabwe, **7** Sally Mugabe Central Hospital, Harare, Zimbabwe

